# Temporal trends in the prescription of biosimilars and the status of switching from original biologics to biosimilars at individual and institutional levels in Japan

**DOI:** 10.1101/2025.06.09.25329307

**Authors:** Minako Matsumoto, Ryosuke Kumazawa, Akiko Ishii-Watabe, Itsuko Horiguchi, Hiroaki Mamiya, Hiroko Shibata, Yoshiro Saito, Motohiko Adomi, Yuta Taniguchi, Jun Komiyama, Ryoko Sakai, Masao Iwagami

## Abstract

**Purpose:** To describe the temporal trends in the prescription of biologics in Japan, with additional analysis focusing on switching from original biologics to biosimilars at the individual and institutional levels.

**Methods:** Using the JMDC claims database from January 2005 to November 2024, we identified patients who received at least one prescription for 17 biologics (original biologics or biosimilars). We elucidated the monthly trends in the proportions of original biologics and biosimilars. We also estimated the proportion of patients receiving original biologics only, those receiving biosimilars only, and those switching from original biologics to biosimilars (and vice versa) during the study period. Finally, we estimated the proportion of medical institutions that started prescribing biosimilars during the study period based on the type of medical institution.

**Results:** Temporal trends in the proportions of original biologics and biosimilars varied widely. In November 2024, the proportion of biosimilar prescriptions was 13.6% for somatropin and 92.5% for filgrastim. At the individual level, the proportion of patients switching from original biologics to biosimilars was low (1.2–14.0%), indicating that switches do not often occur within the same patient, while more recent new users of biologics start biosimilars. At the institutional level, university-related hospitals and clinics were more and less likely, respectively to introduce biosimilars than public and other types of hospitals.

**Conclusion:** Temporal trends in the prescription of biosimilars and switching patterns varied widely by the type of biologics. The type of medical institution should be considered when assessing and promoting the use of biosimilars.

## Introduction

Increasing medical costs has become a global social concern. Prescribed drugs, which account for a considerable part of medical costs, have received attention for potentially reducing its costs [1]. Generic drugs play a pivotal role in reducing drug costs and easing drug budgets [2].

Biologics currently occupy the top positions in drug sales [3]. Biologics are large and complex molecules with structural heterogeneity that have been developed for managing various diseases, including rare diseases [4]. Recently, a group of biologics known as “biosimilars” have been developed to potentially replace high-cost biologics [5]. Biosimilars are biotechnological products that are expected to be comparable to an already approved biotechnological product (referred to as “original biologics”) in terms of quality, efficacy, and safety [6].

Although the global market share of biosimilars is steadily increasing with efforts of governments and industries [7,8], biosimilars have not fully penetrated the biologic market, probably due to concerns of healthcare professionals’ and patients’ regarding their real-world effectiveness and safety [9,10]. A previous systematic review targeting the US and Europe reported that the acceptance of biosimilars varied according to the therapeutic classes [11].

Understanding the trends in the prescription of original biologics and biosimilars is crucial and can be considered the first step towards increasing the use of biosimilars worldwide, including Asia. In addition, researchers and policymakers should understand switches from original biologics to biosimilars at the individual and institutional levels. Replacement of certain original biologics by biosimilars in certain situations or settings, suggesting potential areas for improvement. To our knowledge, no Asian study has systematically assessed the temporal trends and switching patterns from original biologics to biosimilars, covering a variety of biologics [12].

In the present study, we aimed to describe the temporal trends in the prescription of original biologics and biosimilars in Japan, and performed an additional analysis focusing on switching from original biologics to biosimilars at individual and institutional levels.

## Methods

### Data Source

We used data from the JMDC claims database, which has been previously described in detail [13]. Briefly, the JMDC claims database is a large-scale database containing medical claims of large- and medium-sized company employees and their dependent family members aged <75 years. Since 2005, the number of individuals in the JMDC database has increased consistently, reaching a cumulative total of more than 20 million by the end of 2024. The JMDC database includes all the monthly claims for outpatient and inpatient diagnoses and procedures, prescriptions, and dispensations of drugs recorded as the Japanese original drug codes and product names, and the World Health Organization Anatomical Therapeutic Chemical (ATC) classification [14]. In addition, the data included the anonymized IDs of medical institutions, with which we could discern which drug was prescribed by the medical institution, as well as the type of medical institution: clinics (defined in Japan as medical institutions with no or <20 beds for hospitalization), university-related hospitals, public hospitals, and other hospitals (including private hospitals receiving reimbursement from the health insurance system in Japan). We employed the most recent dataset, extracted in December 2024, which included data from January 2005 to November 2024. The data used in this study were anonymized and processed anonymously by JMDC, Inc.

For comparison, we also used the National Database (NDB) Open Data of Health Insurance Claims from April 2022 to March 2023, which is a summary table of the total use of drugs compiled by the government [15] and covers all Japanese citizens except those living with public financial assistance.

This study was approved by the Ethics Committees of the University of Tsukuba, Ibaraki, Japan (approval number 2099) and Meiji Pharmaceutical University, Tokyo, Japan (approval number 202462). The analyses were conducted independently at each study location to verify and obtain similar results.

### Study population

We identified patients receiving at least one prescription of all 17 biologics (for which biosimilars were approved and marketed by 2024) available in Japan during the study period, including somatropin, erythropoietin (including both epoetin alfa and epoetin beta), filgrastim, infliximab, insulin glargine, rituximab, etanercept, trastuzumab, agalsidase beta, bevacizumab, darbepoetin alfa, teriparatide, insulin lispro, adalimumab, insulin aspart, ranibizumab, and pegfilgrastim. For each of the 17 biologics, we created a list of product names (based on at least one prescription record in the JMDC database during the study period) using the ATC classification system, classifying them as original biologics or biosimilars (**Supplementary Table S1**). In the main analysis, we included all these biologics for our analysis. However, for several biologics (somatropin, erythropoietin, insulin glargine, darbepoetin alfa, insulin lispro, and insulin aspart), the original biologics other than the reference product of the biosimilar were approved (**Supplementary Table S1**). Thus, in the sensitivity analysis, we restricted the analysis to biologics and their reference products. In addition, there is one authorized generic (AG) biologic drug for darbepoetin alfa in Japan, which does not have a brand name on its label but is composed of the same drug component as the original biologics [16]. We included this AG biological drug in the biosimilars in the main analysis, but excluded it from our sensitivity analysis.

### Data analysis

After summarizing the demographics (age and sex) of the study population by biologics, we estimated and illustrated the monthly trends in the proportion of prescriptions of original biologics and biosimilars (among the total prescriptions for each biologic) from January 2005 to November 2024. In addition, we compared the statistics (i.e., the number and proportion of biosimilars among the total prescriptions for each biologic) in the JMDC database with those estimated from the NDB Open Data of Health Insurance Claims from April 2022 to March 2023 [15].

Next, at the individual level, we estimated the proportions of (i) patients receiving original biologics only, (ii) those receiving biosimilars only, (iii) those switching from original biologics to biosimilars, (iv) those switching from biosimilars to original biologics, and (v) unknown (because both original biologics and biosimilars were prescribed in the same month, we could not determine which was prescribed earlier from the monthly claims alone) during the study period from January 2005 to November 2024 in the main analysis. Additionally, we restricted the period of the analysis from the time each biosimilar entered the Japanese market to November 2024.

Finally, at the institutional level, among medical institutions with at least one prescription of original biologics, we estimated the proportion of medical institutions starting the prescription of biosimilars during the study period by the type of medical institution (clinics, university-related hospitals, public hospitals, and other hospitals) and 17 biologics. All the analyses were performed using STATA version 17 software (StataCorp, College Station, TX, USA).

## Results

The number of study patients and their demographics varied by biologics, from 102 for agalsidase beta (mean age 38.3 ± 15.4 years, male 56.9%) to 62,038 for insulin glargine (mean age 52.0 ± 12.8 years, male 66.6%) (**Table 1**). In terms of the total number of prescriptions, insulin was most commonly prescribed (insulin glargine, 1,340,426 times; aspart, 1,173,924 times; and lispro, 1,151,886 times), followed by filgrastim (306,931 times). In the sensitivity analyses restricted to biosimilars (excluding AG biologic drugs) and their reference products, the total number of prescriptions decreased, especially for somatropin and darbepoetin alfa.

**Table 1.** Characteristics of studied drugs and study participants.

| Name | ATC code | When the original<br>drug was approved<br>in Japan | When the biosimilar<br>was approved in<br>Japan | Total no. of<br>prescriptions during<br>the study period** | No. of patients<br>with $\geq 1$<br>prescription | Age**, mean<br>± standard<br>deviation | Sex: no. of male<br>patients (%) |
| --- | --- | --- | --- | --- | --- | --- | --- |
| (1) Somatropin | H01AC01 | Nov, 1988 | Jun, 2009 | 277238 | 11264 | 12.4±11.3 | 6557 (58.2) |
| Sensitivity analysis*** | | | | 80791 | 3709 | 11.2±8.9 | 2185 (58.9) |
| (2) Erythropoietin | B03XA01 | Jan, 1990 | Jan, 2010 | 159167 | 20175 | 30.7±27.9 | 8754 (43.4) |
| Sensitivity analysis*** | | | | 112325 | 14344 | 27.1±28.1 | 6312 (44.0) |
| (3) Filgrastim | L03AA02 | Oct, 1991 | Nov, 2012 | 306931 | 25727 | 52.1±15.6 | 12627 (49.1) |
| (4) Infliximab | L04AB02 | Jan, 2002 | Jul, 2014 | 198800 | 10136 | 37.3±15.8 | 6400 (63.1) |
| (5) Insulin glargine | A10AE04 | Oct, 2003 | Dec, 2014 | 1340426 | 62038 | 52.0±12.8 | 41289 (66.6) |
| Sensitivity analysis*** | | | | 1017736 | 51381 | 52.3±12.6 | 34487 (67.1) |
| (6) Rituximab | L01FA01 | Jun, 2001 | Sep, 2017 | 87361 | 8252 | 51.5±16.2 | 4868 (59.0) |
| (7) Etanercept | L04AB01 | Jan, 2005 | Jan, 2018 | 192029 | 7521 | 49.2±12.0 | 1203 (16.0) |
| (8) Trastuzumab | L01FD01 | Apr, 2001 | Mar, 2018 | 227537 | 9136 | 52.4±9.2 | 523 (5.7) |
| (9) Agalsidase beta | A16AB04 | Jan, 2004 | Sep, 2018 | 11869 | 102 | 38.3±15.4 | 58 (56.9) |
| (10) Bevacizumab | L01FG01 | Apr, 2007 | Jun, 2019 | 268449 | 14261 | 55.0±10.6 | 5691 (39.9) |
| (11) Darbepoetin alfa | B03XA02 | Apr, 2007 | Sep, 2019 | 131241 | 13083 | 55.2±12.6 | 8480 (64.8) |
| Sensitivity analysis*** | | | | 81572 | 8491 | 55.0±12.6 | 5440 (64.1) |
| (12) Teriparatide | H05AA02 | Jul, 2010 | Sep, 2019 | 74155 | 5345 | 62.3±8.6 | 938 (17.6) |
| (13) Insulin lispro | A10AB04 | Aug, 2001 | Mar, 2020 | 1151886 | 55511 | 49.8±13.9 | 32012 (57.7) |
|  | A10AC04 |  |  |  |  |  |  |
|  | A10AD04 |  |  |  |  |  |  |
| Sensitivity analysis*** | | | | 932485 | 48537 | 49.6±14.0 | 27663 (57.0) |
| (14) Adalimumab | L04AB04 | Apr, 2008 | Jun, 2020 | 196998 | 10558 | 40.3±15.1 | 5655 (53.6) |
| (15) Insulin aspart | A10AB05<br>A10AD05 | Apr, 2008 | Mar, 2021 | 1173924 | 49859 | 49.7±14.7 | 28693 (57.6) |
| Sensitivity analysis*** |  |  |  | 971582 | 44243 | 49.3±14.8 | 25123 (56.8) |
| (16) Ranibizumab | S01LA04 | Jan, 2009 | Sep, 2021 | 23925 | 8160 | 56.0±12.8 | 4846 (59.4) |
| (17) Pegfilgrastim | L03AA13 | Sep, 2014 | Sep, 2023 | 93019 | 19921 | 53.0±11.0 | 5367 (26.9) |

\*Age at the time of first prescription in the JMDC claims database.
\*\*From January 2005 to May 2024
\*\*\*While all the original biologics or biosimilars available in Japan were included in the main analysis, the sensitivity analysis made the changes below (corresponding to Supplementary Table S1):
- For somatropin, we restricted to Genotropin vs. Somatropin BS. - For erythropoietin, we restricted to Espo vs. Epoetin Alfa BS (Epoetin Kappa). - For insulin glargine, we restricted to Lantus (not including Lantus XR) vs. Insulin Glargine BS. - For darbepoetin alfa, we restricted to Nesp vs. Darbepoetin Alfa BS (not including Darbepoetin Alfa authorized generic). - For insulin lispro, we restricted to Humalog (not including Humalog Mix and Humalog N) vs. Insulin Lispro BS. - For insulin aspart, we restricted to NovoRapid (not including NovoRapid Mix) vs. Insulin Aspart BS.

The monthly trends in the proportion of original biologics and biosimilars varied widely among biologics (**Fig 1**). Some biologics, such as filgrastim and trastuzumab, demonstrated a steep increase in the proportion of biosimilars since their launch, whereas biologics, such as somatropin and infliximab, demonstrated a slow increase. Darbepoetin alpha and insulin lispro initially demonstrated a steep increase, which became nearly flat. In November 2024, the proportion of biosimilar prescriptions (among all prescriptions of biologics) ranged from 13.6% for somatropin to 92.5% for filgrastim.

**Fig 1.**
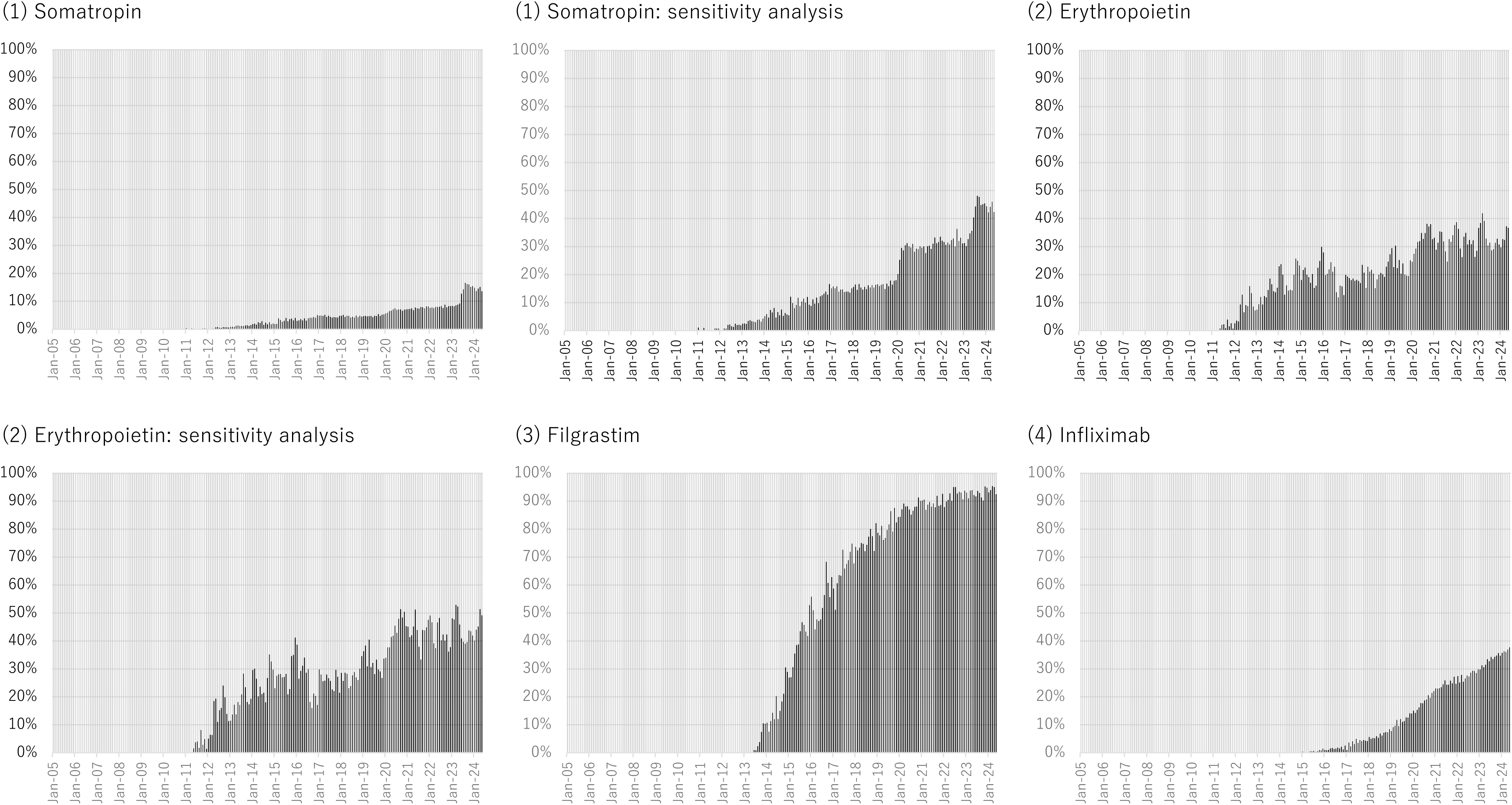
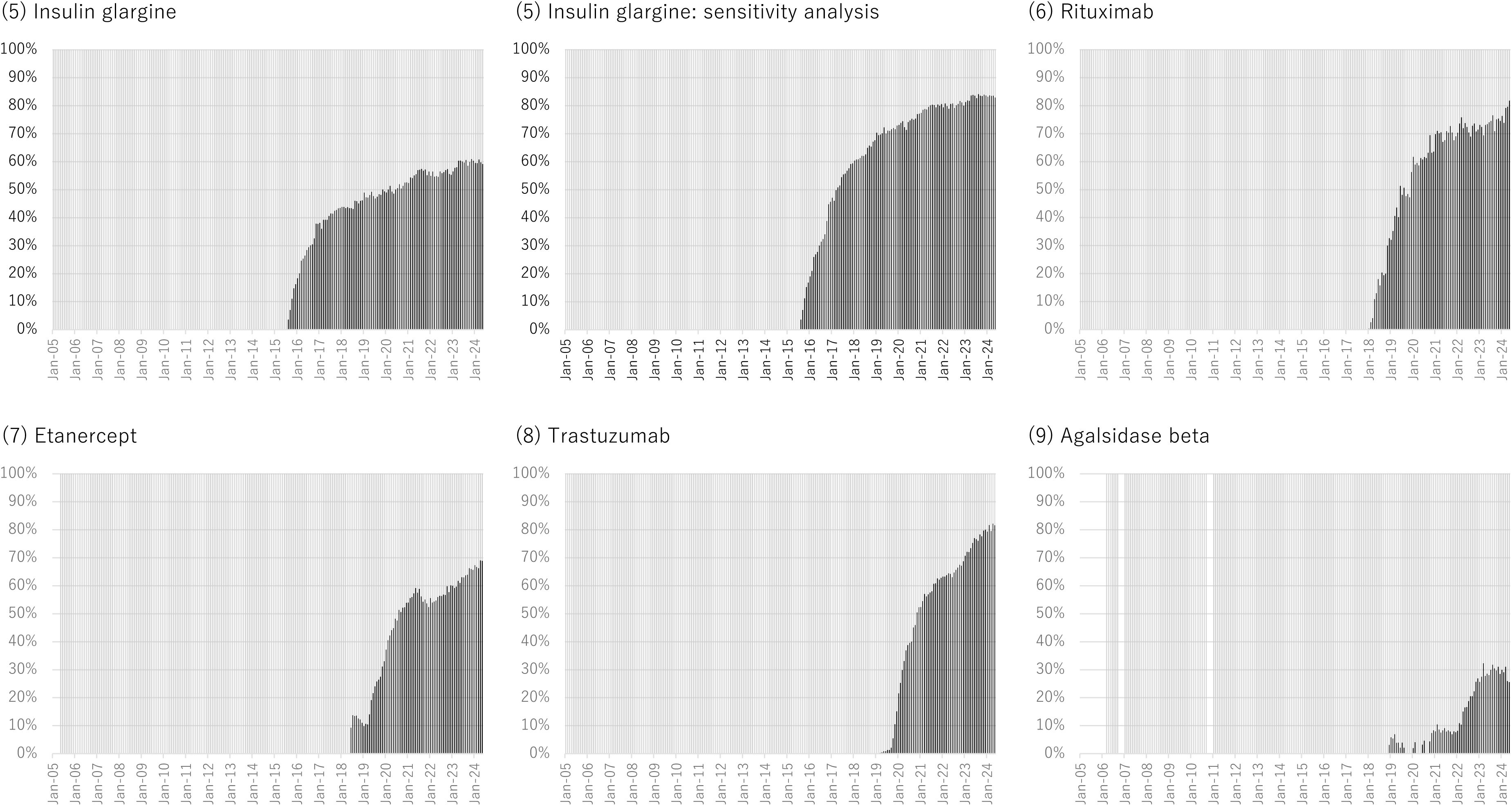
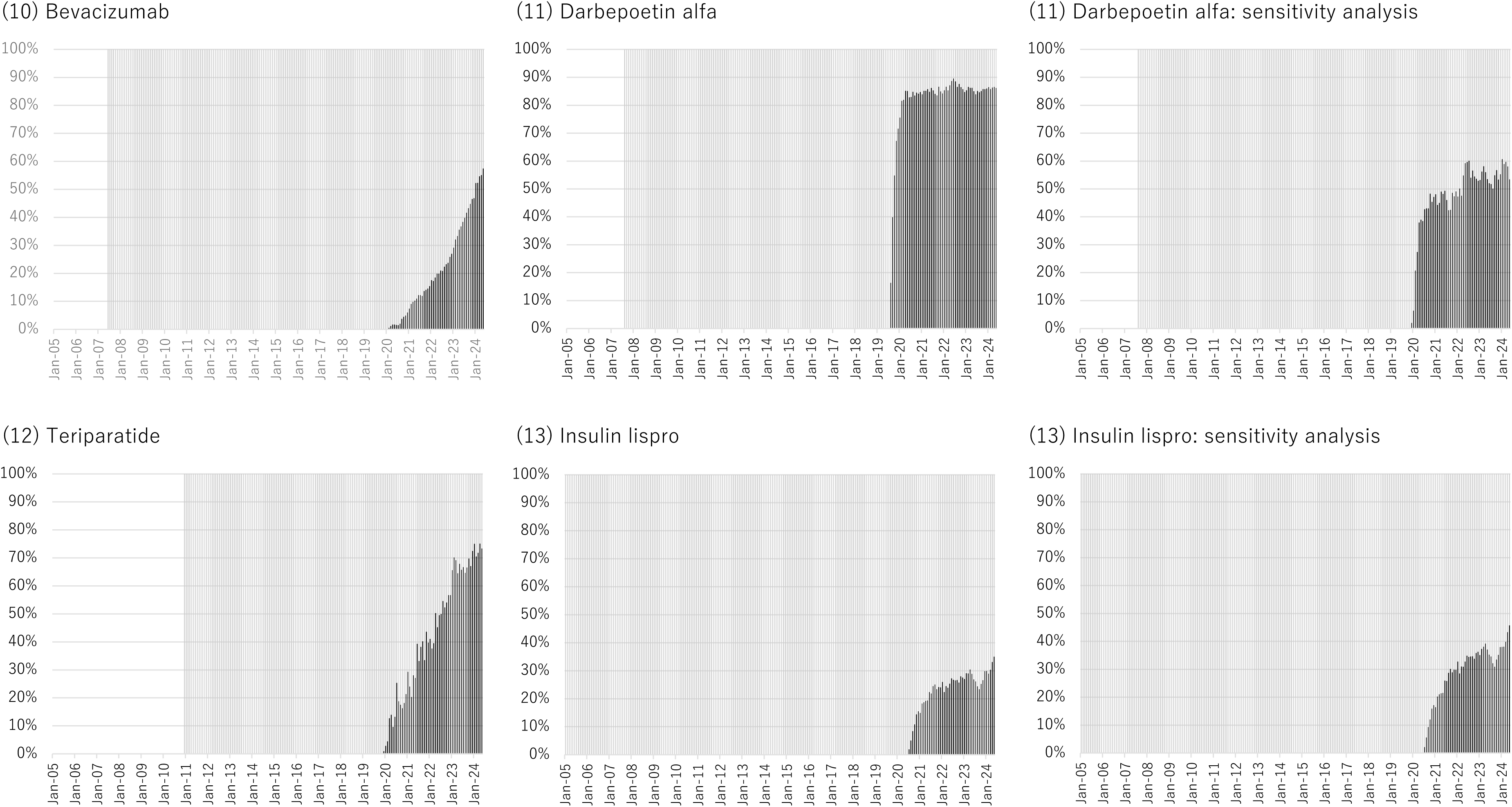
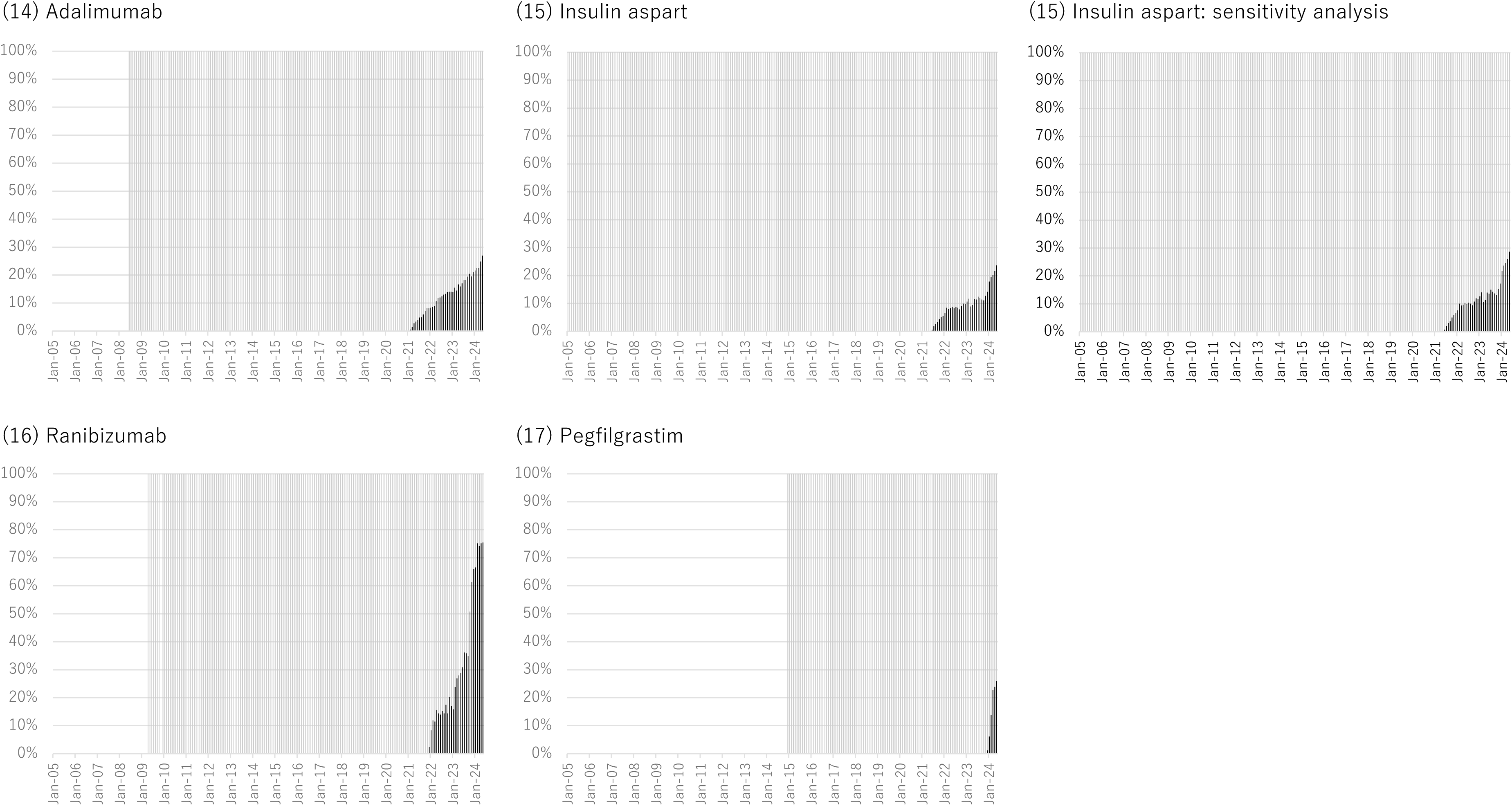
Monthly trend in the proportion of prescriptions of original biologics (colored gray) or biosimilars (colored black) among the total prescriptions for each biologic during the study period. Note: Although all original biologics or biosimilars available in Japan were included in the main analysis, the sensitivity analysis made the following changes (Supplementary Table S1):

- For somatropin, we considered Genotropin vs. Somatropin BS.
- For erythropoietin, we considered Espo vs. Epoetin Alfa BS (Epoetin Kappa).
- For insulin glargine, we considered Lantus (not including Lantus XR) vs. Insulin Glargine BS.
- For darbepoetin alfa, we considered Nesp vs. Darbepoetin Alfa BS (not including Darbepoetin Alfa authorized generic).
- For insulin lispro, we considered Humalog (not including Humalog Mix and Humalog N) vs. Insulin Lispro BS.
- For insulin aspart, we considered NovoRapid (not including NovoRapid Mix) vs. Insulin Aspart BS.

The statistics in the JMDC claims database and the NDB Open Data from April 2022 to March 2023 were mostly similar (**Supplementary Table S2**), suggesting the generalizability of the findings in the present study.

Supplementary Table S3. and **Fig 2** illustrate the distribution of patients receiving original biologics only, biosimilars only, and those switching from original biologics to biosimilars, or vice versa. The proportion of patients switching from the original biologics to biosimilars was generally low, varying from 1.2% for erythropoietin to 14.0% for etanercept. The proportion of patients receiving biosimilars only was much higher than that of the patients switching for all biologics, implying that switches do not often occur within the same patient, whereas more recent new users of biologics start with biosimilars. The proportion of patients receiving biosimilars only was the highest for filgrastim (74.4%), followed by darbepoetin alfa (45.4%) and insulin glargine (43.9%). In the additional analysis, restricting the analysis period from when each biologic was launched (to November 2024), the proportion of switchers from the original biologics to biosimilars did not change significantly and remained low, whereas the proportion of patients receiving only biosimilars tended to increase (**Supplementary Table S3**).

**Fig 2.**
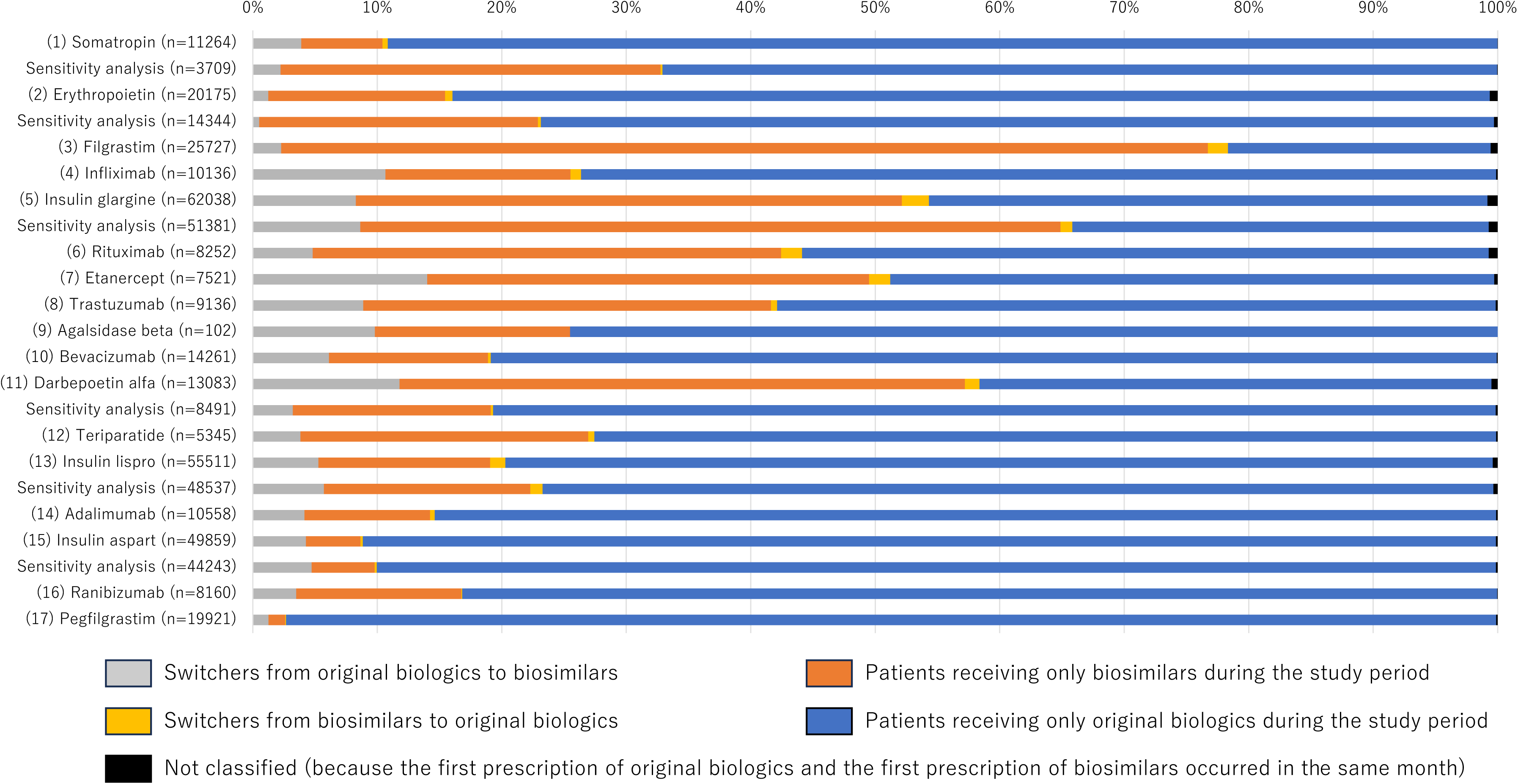
Distribution of patients receiving only original biologics or biosimilars during the study period or switchers. Note: For switchers, only the first switch was assessed and counted (i.e., some patients switched twice or more). Note: Although all original biologics or biosimilars available in Japan were included in the main analysis, the sensitivity analysis made the following changes (Supplementary Table S1):

- For somatropin, we considered Genotropin vs. Somatropin BS.
- For erythropoietin, we considered Espo vs. Epoetin Alfa BS (Epoetin Kappa).
- For insulin glargine, we considered Lantus (not including Lantus XR) vs. Insulin Glargine BS.
- For darbepoetin alfa, we considered Nesp vs. Darbepoetin Alfa BS (not including Darbepoetin Alfa authorized generic).
- For insulin lispro, we considered Humalog (not including Humalog Mix and Humalog N) vs. Insulin Lispro BS.
- For insulin aspart, we considered NovoRapid (not including NovoRapid Mix) vs. Insulin Aspart BS.

Finally, at the institutional level, for most biologics, the proportion of medical institutions introducing biosimilars during the study period (among those once prescribing original biologics as the denominator) was the highest in university-related hospitals, followed by public hospitals, other hospitals, and clinics (**Fig 3**). However, insulin glargine and darbepoetin alfa were relatively commonly prescribed in all types of medical institutions compared with other biologics.

**Fig 3.**
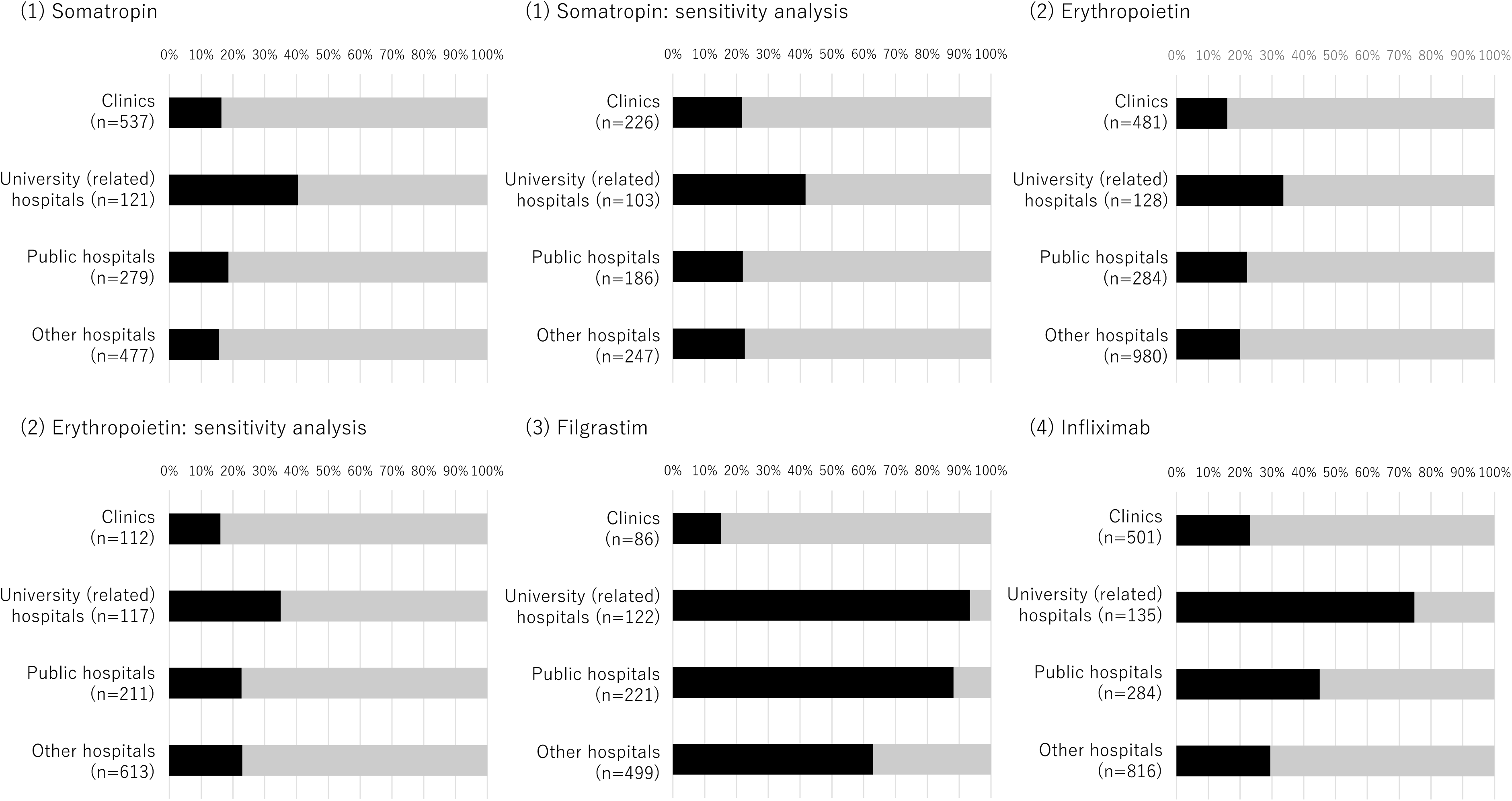
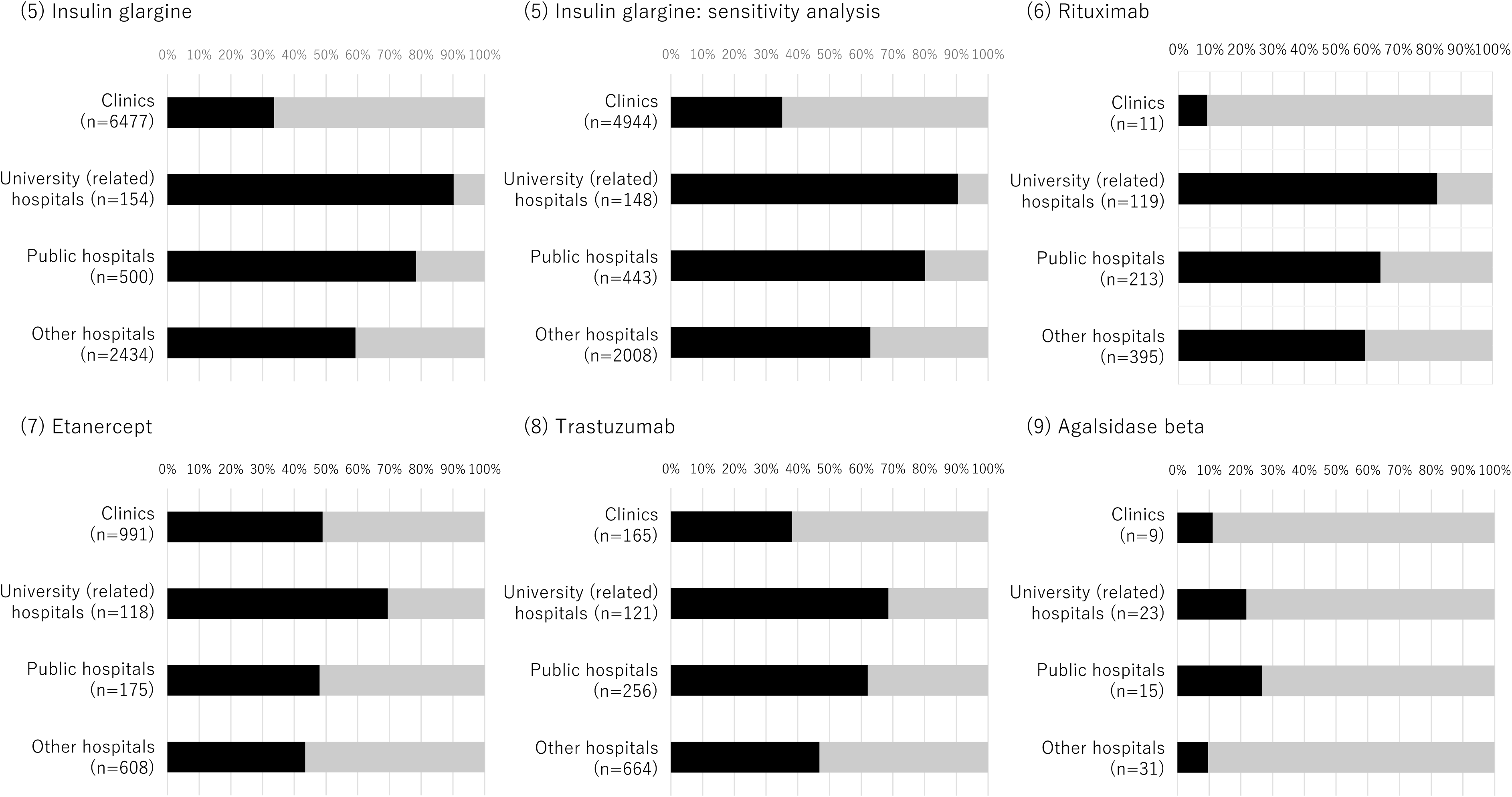
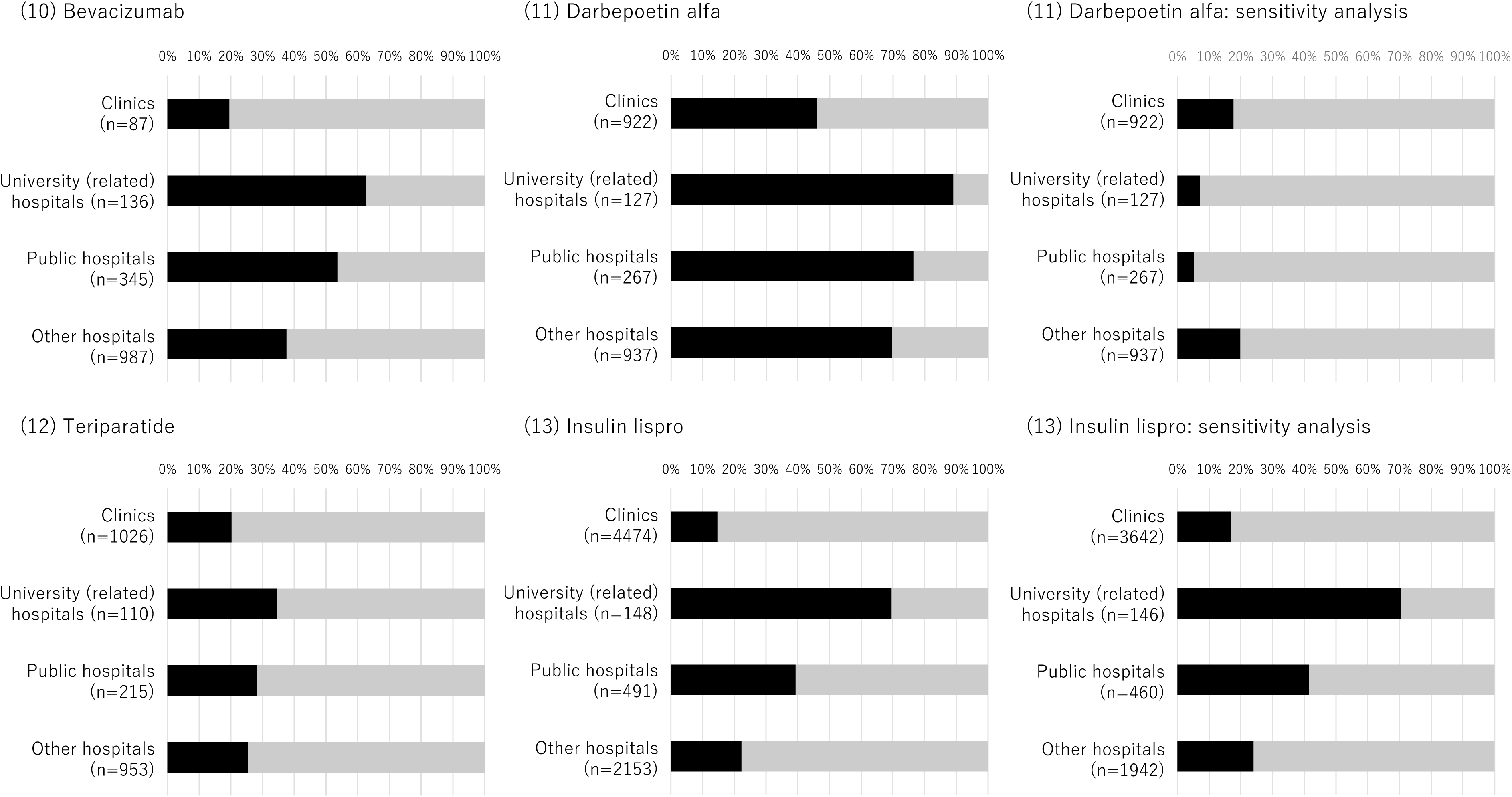
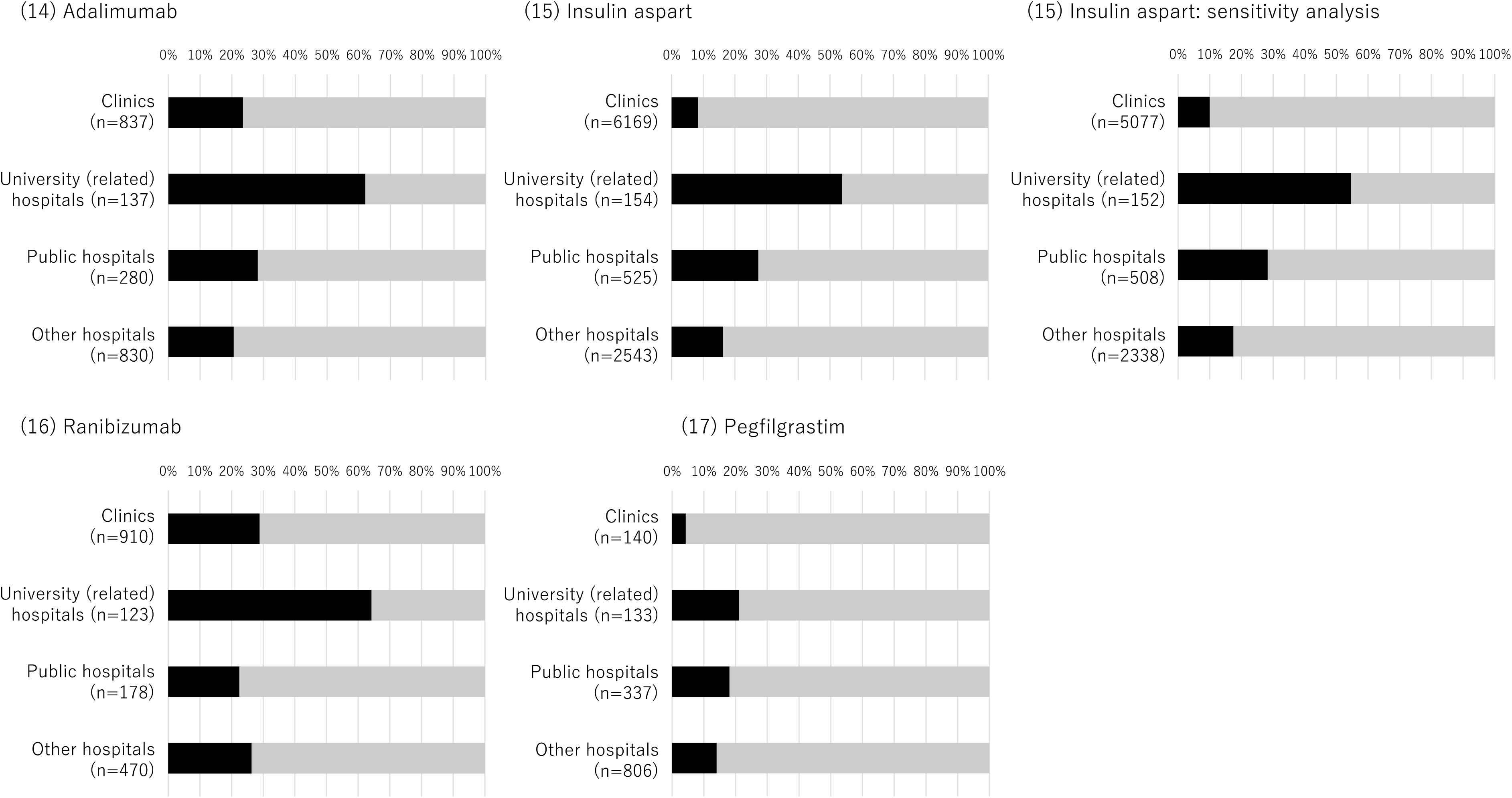
Distribution of medical institutions prescribing only original biologics (colored gray) or original biologics and biosimilars (colored black) during the study period. Note: Although all original biologics or biosimilars available in Japan were included in the main analysis, the sensitivity analysis made the following changes (Supplementary Table S1):

- For somatropin, we considered Genotropin vs. Somatropin BS.
- For erythropoietin, we considered Espo vs. Epoetin Alfa BS (Epoetin Kappa).
- For insulin glargine, we considered Lantus (not including Lantus XR) vs. Insulin Glargine BS.
- For darbepoetin alfa, we considered Nesp vs. Darbepoetin Alfa BS (not including Darbepoetin Alfa authorized generic).
- For insulin lispro, we considered Humalog (not including Humalog Mix and Humalog N) vs. Insulin Lispro BS.
- For insulin aspart, we considered NovoRapid (not including NovoRapid Mix) vs. Insulin Aspart BS.

## Discussion

This study comprehensively investigated the temporal trends in the prescription of biosimilars, with an additional analysis focusing on switching from the original biologics to biosimilars at the individual and institutional levels, using the JMDC claims database for company employees and their dependent family members. The introduction of biosimilars varied widely by the type of biologics as well as by the type of medical institution. Switching within the same individual was uncommon, whereas more recent new biologics users started using biosimilars only. At the institutional level, university-related hospitals and clinics were more and less likely, respectively, to introduce biosimilars than public and other types of hospitals.

The most frequently prescribed biologics were insulin (glargine, aspart, and lispro) merely because diabetes is a common disease, with an estimated 11 million patients in Japan in 2021 [17]. The number of prescriptions of insulin and filgrastim was proportional to the number of patients, while some biologics, such as erythropoietin and pegfilgrastim, were not, suggesting that the frequency and duration of use in individuals vary according to the biologics and related underlying diseases. Restricting our analysis to biosimilars (excluding AG biological drugs) and their reference products, the levels of somatropin and darbepoetin alfa decreased. This suggests that most of the patients treated with somatropin used the original biologics, which were not the reference products, whereas most of the patients treated with darbepoetin alfa used AG biologic drugs.

The monthly trends in the proportion of biosimilars varied widely according to the biologics. The use of biosimilars filgrastim, rituximab, trastuzumab, ranibizumab, and teriparatide demonstrated rapid increase during the study period. We did not find consistent characteristics that were obviously different between biologics with and without a rapid increase in the proportion of biosimilars, such as the timing of approval and underlying diseases. For example, biosimilars used in cancer treatment, such as filgrastim, rituximab, and trastuzumab, have demonstrated a rapid increase in use; however, bevacizumab (also used in cancer treatment) did not demonstrate a similar trend.

We found some temporal stagnation or a decrease in the proportion of several biologics, such as etanercept, darbepoetin alfa, and filgrastim. One possible explanation is that some practitioners may have stopped using biosimilars owing to a shortage of biosimilars in Japan. Drug shortages occurred for etanercept in 2018 and 2021, darbepoetin alfa in 2020, filgrastim in 2020 and 2022, and pegfilgrastim in 2024. One of the major reasons for drug shortages is that the demand exceeds expectations. A stable supply of biosimilars is necessary to promote their use.

The proportion of switchers from original biologics to biosimilars was generally small. One obvious reason for the low switching proportion within the same individual is that switching from an original biologic to a biosimilar over the course of treatment was not recommended in Japan until 2022 to ensure the traceability of the products as per domestic guidelines. In addition, a previous study reported that some physicians [18] and patients [19] may be concerned about the safety profiles of biosimilars, especially when they switch from the original biologics to biosimilars, which can lead to negative expectations and nocebo effects [20]. A better understanding of biosimilars could help encourage their acceptance by prescribers and patients.

The introduction of biosimilars varies according to the type of medical institution. The proportion of clinics that introduced biosimilars was lower than that of the hospitals. University-related hospitals demonstrated the highest proportion of biosimilars. We speculate that hospitals (especially university-related hospitals) may be able to reduce medical costs by introducing biosimilars, or may be more likely to be influenced by the government policy of promoting biosimilars than clinics. As opposed to physicians in hospitals who follow the hospital formulary, physicians in clinics can select any drug based on the preferences of the physician and patient. To increase biosimilar usage, we need to further explore the factors affecting healthcare providers’ and patients’ acceptance of switching to biosimilars.

This study has certain limitations. First, the database comprised large and medium-sized company employees and their family members; thus, the population of this study is expected to be younger and more affluent than the average Japanese population. However, most biologics are prescribed for patients aged <65 years, and we confirmed that the statistics in the JMDC claims database was generally similar to that obtained from the NDB Open data, which are nationally representative statistics. Second, the present findings from Japan would be informative to, but may not be directly applicable to, other countries because biosimilar usage and switching patterns are affected by various factors such as healthcare policies and reimbursement systems. Finally, although the medical institution IDs and types of medical institutions were variable information in the JMDC claims database, we were unable to obtain other information, such as the area (region) of the medical institutions and the socioeconomic status of the area.

## Conclusions

Trends in biologics utilization varied widely between biologics and medical institutions. Data capturing the current use of original biologics and biosimilars in a real-world setting provided the characteristics of biologics, which may contribute to the development of targeted interventions, thus promoting efficient and effective use of biosimilars in the future.

## Data Availability

We obtained data from JMDC Inc. and did not obtain permission to share these data with other parties. Researchers who meet the access criteria can acquire de-identified participant data from JMDC Inc. (https://www.jmdc.co.jp/en/).

## Acknowledgment

The authors would like to thank the contribution of Hiroyuki Sakamaki (Kanagawa University of Human Services, as well as Pharma Policy Planning P-cubed) to advice on the classification of original biologics and biosimilars.

## Author Contributions

MI planned the study and obtained the data. MI, YT, JK, RK, and RS conducted the analysis and created the tables and figures. MM wrote the first draft of the manuscript. AIW, IH, HM, HS, YS, and MA provided critical comments for improving the methods and discussion. All authors have read the final version of the manuscript and have agreed to its submission.

## References

1. Annual Health, Labour and Welfare Report 2021. https://www.mhlw.go.jp/english/wp/wp-hw14/dl/02e.pdf

2. Kleinstauber M. The Role of Generic Medicines in Reducing Healthcare Costs. J Adv Pract Nurs. 2024;09:4L5.

3. Rieger C, Dean JA, Hall L, et al. Barriers and Enablers Affecting the Uptake of Biosimilar Medicines Viewed Through the Lens of Actor Network Theory: A Systematic Review. BioDrugs. 2024;38:541–555.

4. Liu Y, Yang M, Garg V, et al. Economic Impact of Non-Medical Switching from Originator Biologics to Biosimilars: A Systematic Literature Review. Adv Ther. 2019;36:1851–1877.

5. Mirjalili SZ, Sabourian R, Sadeghalvad M, et al. Therapeutic applications of biosimilar monoclonal antibodies: Systematic review of the efficacy, safety, and immunogenicity in autoimmune disorders. Int Immunopharmacol. 2021;101:108305.

6. Ishii-Watabe A, Kuwabara T. Biosimilarity assessment of biosimilar therapeutic monoclonal antibodies. Drug Metab Pharmacokinet. 2019;34:64–70.

7. The Impact of Biosimilar Competition. Available from: https://www.medicinesforeurope.com/wp-content/uploads/2016/03/IMS-Impact-of-Biosimilar-Competition-2015.pdf

8. Kim Y, Kwon H, Godman B, et al. Uptake of Biosimilar In fl iximab in the UK, France, Japan, and KoreaL: Budget Savings or Market Expansion Across CountriesL? Front Pharmacol. 2020;11:1–12.

9. Sarnola K, Merikoski M, Jyrkkä J, et al. Physicians’ perceptions of the uptake of biosimilarsL: a systematic review. BMJ Open. 2020;10:e034183.

10. Wu Q, Wang Z, Wang X, et al. Patients’ Perceptions of BiosimilarsL: A Systematic Review. BioDrugs. 2023;37:829–841.

11. Leonard E. Factors Affecting Health Care Provider Knowledge and Acceptance of Biosimilar Medicines: A Systematic Review. J Manag Care Spec Pharm. 2019;25:102–112.

12. Roth JA, Rahshenas M, Nowacki G, et al. A descriptive analysis of real-world oncology biosimilar use in Japan. Futur Oncol. 2024;20:1837–1850.

13. Nagai K, Tanaka T, Kodaira N, et al. Data resource profile: JMDC claims databases sourced from Medical Institutions. J Gen Fam Med. 2020;21:211–218.

14. WHO. Guideline for ATC classification and DDD assignment 2024.

15. Yasunaga H. Updated Information on NDB. Ann Clin Epidemiol. 2024;6:73–76.

16. Gulsen Oner Z, Polli JE. Authorized Generic Drugs: an Overview. AAPS PharmSciTech. 2018;19:2450–2458.

17. International Diabetes Federation. Available from: https://diabetesatlas.org/data-by-location/country/japan/

18. Cohen H, Beydoun D, Chien D, et al. Awareness, Knowledge, and Perceptions of Biosimilars Among Specialty Physicians. Adv Ther. 2017;33:2160–2172.

19. Jacobs I, Singh E, Sewell KL. Patient attitudes and understanding about biosimilars: An international cross-sectional survey. Patient Prefer Adherence. 2016;10:937–948.

20. Rezk MF, Pieper B. Treatment Outcomes with Biosimilars: Be Aware of the Nocebo Effect. Rheumatol Ther. 2017;4:209–218.

