## Supplementary Tables for "Temporal trends in the prescription of biosimilars and the status of switching from original biologics to biosimilars at individual and institutional levels in Japan"

**Supplementary Table S1. List of product names and classifications**

| Name | ATC code | Original biologic | Biosimilar |
| --- | --- | --- | --- |
| 1. Somatropin | H01AC01 | Genotropin* | Somatropin BS |
|  |  | Growject | – |
|  |  | Saizen | – |
|  |  | Norditropin | – |
|  |  | Humatrope | – |
| 2. Erythropoietin*** | B03XA01 | Espo* | Epoetin Alfa BS  (Epoetin Kappa) |
|  |  | Epogin |  |
| 3. Filgrastim | L03AA02 | Gran* | Filgrastim BS |
| 4. Infliximab | L04AB02 | Remicade* | Infliximab BS |
| 5. Insulin glargine | A10AE04 | Lantus* | Insulin Glargine BS |
|  |  | Lantus XR | – |
| 6. Rituximab | L01FA01 | Rituxan* | Rituximab BS |
| 7. Etanercept | L04AB01 | Enbrel* | Etanercept BS |
| 8. Trastuzumab | L01FD01 | Herceptin* | Trastuzumab BS |
| 9. Agalsidase beta | A16AB04 | Fabrazyme* | Agalsidase Beta BS |
| 10. Bevacizumab | L01FG01 | Avastin* | Bevacizumab BS |
| 11. Darbepoetin alfa | B03XA02 | Nesp* | Darbepoetin Alfa BS |
|  |  |  | Darbepoetin Alfa** |
| 12. Teriparatide | H05AA02 | Forteo* | Teriparatide BS |
| 13. Insulin lispro | A10AB04  A10AC04  A10AD04 | Humalog* | Insulin Lispro BS |
|  |  | Humalog Mix | – |
|  |  | Humalog N | – |
|  |  | Lyumjev | – |
| 14. Adalimumab | L04AB04 | Humira* | Adalimumab BS |
| 15. Insulin aspart | A10AB05  A10AD05 | NovoRapid* | Insulin Aspart BS |
|  |  | NovoRapid Mix | – |
|  |  | Fiasp | – |
| 16. Ranibizumab | S01LA04 | Lucentis* | Ranibizumab BS |
| 17. Pegfilgrastim | L03AA13 | G-lasta* | Pegfilgrastim BS |

* Reference product of biosimilar.

** Authorized generic (AG) biologic drug.

***Different from other biologics, “Erythropoietin” is not a generic name but used here because multiple active ingredients exist for original biologics, including epoetin alfa (Espo) and epoetin beta (Epogin).

**Supplementary Table S2. Comparison of number of prescriptions and propotion of biosimilars between the JMDC claims database and the NDB Open Data from April 2022 to March 2023**

| Name | JMDC claims database from April 2022 to March 2023 | | NDB Open Data from April 2022 to March 2023 | |
| --- | --- | --- | --- | --- |
|  | Total no. of prescriptions for both original biologics and biosimilars | No. of prescriptions for biosimilars (%) | Total no. of prescriptions for both original biologics and biosimilars | Total no. of prescriptions for both original biologics and biosimilars |
| (1) Somatropin | 34,931 | 2,859 (8.2) | 1087,245 | 84,386 (7.8) |
| Sensitivity analysis* | 8,943 | 2,859 (32.0) | 285,465 | 84,386 (29.6) |
| (2) Erythropoietin | 16,083 | 5,278 (32.8) | 337,188 | 121,294 (36.0) |
| Sensitivity analysis* | 12,140 | 5,278 (43.5) | 186,132 | 121,294 (65.2) |
| (3) Filgrastim | 39,343 | 36,552 (92.9) | 907,989 | 861,176 (94.8) |
| (4) Infliximab | 25,590 | 7,349 (28.7) | 937,777 | 272,924 (29.1) |
| (5) Insulin glargine | 162,849 | 91,755 (56.3) | 6070,213 | 3267,121 (53.8) |
| Sensitivity analysis* | 113,707 | 91,755 (80.7) | 4446,768 | 3267,121 (73.5) |
| (6) Rituximab | 11,134 | 7,986 (71.7) | 357,789 | 282,047 (78.8) |
| (7) Etanercept | 23,607 | 13,654 (57.8) | 1925,756 | 898,041 (46.6) |
| (8) Trastuzumab | 29,597 | 19,915 (67.3) | 918,232 | 602,321 (65.6) |
| (9) Agalsidase beta | 1,808 | 411 (22.7) | 45,713 | 8,063 (17.6) |
| (10) Bevacizumab | 37,689 | 9,426 (25.0) | 1577,373 | 425,443 (27.0) |
| (11) Darbepoetin alfa | 14,436 | 12,550 (86.9) | 1518,781 | 1198,118 (78.9) |
| Sensitivity analysis* | 4,293 | 2,407 (56.1) | 522,992 | 201,751 (38.6) |
| (12) Teriparatide | 9,071 | 5,082 (56.0) | 754,299 | 329,326 (43.7) |
| (13) Insulin lispro | 156,459 | 42,161 (26.9) | 9079,238 | 1824,060 (20.1) |
| Sensitivity analysis* | 120,711 | 42,161 (34.9) | 6552,217 | 1824,060 (27.8) |
| (14) Adalimumab | 29,496 | 3,926 (13.3) | 1090,901 | 136,087 (12.5) |
| (15) Insulin aspart | 132,168 | 12,139 (9.2) | 8663,964 | 535,289 (6.2) |
| Sensitivity analysis* | 110,409 | 12,139 (11.0) | 6693,072 | 535,289 (8.0) |
| (16) Ranibizumab | 3,464 | 603 (17.4) | 168,422 | 26,819 (15.9) |
| (17) Pegfilgrastim | 16,297 | 0 (0.0) | 306,436 | 0 (0.0) |

NDB, National Database.

*While all the original biologics or biosimilars available in Japan were included in the main analysis, the sensitivity analysis made the changes below (corresponding to Supplementary Table S1):

- For somatropin, we considered Genotropin vs. Somatropin BS.
- For erythropoietin, we considered Espo vs. Epoetin Alfa BS (Epoetin Kappa).
- For insulin glargine, we considered Lantus (not including Lantus XR) vs. Insulin Glargine BS.
- For darbepoetin alfa, we considered Nesp vs. Darbepoetin Alfa BS (not including Darbepoetin Alfa authorized generic).
- For insulin lispro, we considered Humalog (not including Humalog Mix and Humalog N) vs. Insulin Lispro BS.
- For insulin aspart, we considered NovoRapid (not including NovoRapid Mix) vs. Insulin Aspart BS.

**Supplementary Table S3. Distribution of patients using only original biologics or biosimilars or switchers during the study period**

| Drug | Main analysis from Jan, 2005 to May, 2024 | | | | | | Additional analysis restricting to patients using drugs only during the period from when each biosimilar was approved to May, 2024 | | | | | | |
| --- | --- | --- | --- | --- | --- | --- | --- | --- | --- | --- | --- | --- | --- |
|  | Switchers from original biologics to biosimilars | Patients using only biosimilars | Switchers from biosimilars to original biologics | Patients using only original biologics | Not classified* | Total no. of patients | When each biosimilar was approved | Switchers from original biologics to biosimilars | Patients using only biosimilars | Switchers from biosimilars to original biologics | Patients using only original biologics | Not classified* | Total no. of patients |
| (1) Somatropin | 440  (3.9%) | 734  (6.5%) | 47  (0.4%) | 10,042  (89.2%) | 1  (<0.1%) | 11,264  (100%) | Jun, 2009 | 439  (4.0%) | 734  (6.6%) | 47  (0.4%) | 9,835  (89.0%) | 1  (<0.1%) | 11,056  (100%) |
| Sensitivity analysis** | 83  (2.2%) | 1,132  (30.5%) | 6  (0.2%) | 2,487  (67.1%) | 1  (<0.1%) | 3,709  (100%) |  | 82  (2.3%) | 1,132  (31.2%) | 6  (0.2%) | 2,404  (66.3%) | 1  (<0.1%) | 3,625  (100%) |
| (2) Erythropoietin | 252  (1.2%) | 2,864  (14.2%) | 122  (0.6%) | 16,812  (83.3%) | 125  (0.6%) | 20,175  (100%) | Jan, 2010 | 234  (1.2%) | 2,864  (14.7%) | 122  (0.6%) | 16,102  (82.8%) | 125  (0.6%) | 19,447  (100%) |
| Sensitivity analysis** | 74  (0.5%) | 3,211  (22.4%) | 36  (0.3%) | 10,981  (76.6%) | 42  (0.3%) | 14,344  (100%) |  | 68  (0.5%) | 3,211  (23.2%) | 36  (0.3%) | 10,509  (75.8%) | 42  (0.3%) | 13,866  (100%) |
| (3) Filgrastim | 590  (2.3%) | 19,151  (74.4%) | 415  (1.6%) | 5,427  (21.1%) | 144  (0.6%) | 25,727  (100%) | Nov, 2012 | 558  (2.3%) | 19,151  (78.5%) | 415  (1.7%) | 4,129  (16.9%) | 144  (0.6%) | 24,397  (100%) |
| (4) Infliximab | 1,080  (10.7%) | 1,507  (14.9%) | 86  (0.8%) | 7,451  (73.5%) | 12  (0.1%) | 10,136  (100%) | Jul, 2014 | 918  (10.7%) | 1,507  (17.5%) | 86  (1.0%) | 6,090  (70.7%) | 12  (0.1%) | 8,613  (100%) |
| (5) Insulin glargine | 5,142  (8.3%) | 27,208  (43.9%) | 1,350  (2.2%) | 27,838  (44.9%) | 500  (0.8%) | 62,038  (100%) | Dec, 2014 | 3,603  (6.7%) | 27,208  (50.4%) | 1,350  (2.5%) | 21,361  (39.5%) | 500  (0.9%) | 54,022  (100%) |
| Sensitivity analysis** | 4,440  (8.6%) | 28,903  (56.3%) | 489  (1.0%) | 17,181  (33.4%) | 368  (0.7%) | 51,381 (100%) |  | 2,901  (6.7%) | 28,903  (66.7%) | 489  (1.1%) | 10,704  (24.7%) | 368  (0.9%) | 43,365  (100%) |
| (6) Rituximab | 398  (4.8%) | 3,105  (37.6%) | 138  (1.7%) | 4,552  (55.2%) | 59  (0.7%) | 8,252  (100%) | Sep, 2017 | 279  (4.5%) | 3,105  (49.5%) | 138  (2.2%) | 2,693  (42.9%) | 59  (0.9%) | 6,274  (100%) |
| (7) Etanercept | 1,053  (14.0%) | 2,671  (35.5%) | 128  (1.7%) | 3,649  (48.5%) | 20  (0.3%) | 7,521  (100%) | Jan, 2018 | 529  (10.5%) | 2,671  (53.0%) | 128  (2.5%) | 1,695  (33.6%) | 20  (0.4%) | 5,043  (100%) |
| (8) Trastuzumab | 811  (8.9%) | 2,992  (32.7%) | 45  (0.5%) | 5,275  (57.7%) | 13  (0.1%) | 9,136  (100%) | Mar, 2018 | 687  (11.1%) | 2,992  (48.3%) | 45  (0.7%) | 2,463  (39.7%) | 13  (0.2%) | 6,200  (100%) |
| (9) Agalsidase beta | 10  (9.8%) | 16  (15.7%) | 0  (0%) | 76  (74.5%) | 0  (0%) | 102  (100%) | Sep, 2018 | 4  (5.7%) | 16  (22.9%) | 0  (0%) | 50  (71.4%) | 0  (0%) | 70  (100%) |
| (10) Bevacizumab | 873  (6.1%) | 1,823  (12.8%) | 31  (0.2%) | 11,525  (80.8%) | 9  (0.1%) | 14,261  (100%) | Jun, 2019 | 747  (8.8%) | 1,823  (21.5%) | 31  (0.4%) | 5,881  (69.3%) | 9  (0.1%) | 8,491  (100%) |
| (11) Darbepoetin alfa | 1,542  (11.8%) | 5,942  (45.4%) | 153  (1.2%) | 5,382  (41.1%) | 64  (0.5%) | 13,083  (100%) | Sep, 2019 | 216  (3.1%) | 5,927  (85.8%) | 153  (2.2%) | 557  (8.1%) | 57  (0.8%) | 6,910  (100%) |
| Sensitivity analysis** | 273  (3.2%) | 1,350  (5.9%) | 16  (0.2%) | 6,839  (80.5%) | 13  (0.2%) | 8,491  (100%) |  | 65  (2.8%) | 1,350  (57.9%) | 16  (0.7%) | 889  (38.1%) | 13  (0.6%) | 2,333  (100%) |
| (12) Teriparatide | 205  (3.8%) | 1,236  (23.1%) | 26  (0.5%) | 3,872  (72.4%) | 6  (0.1%) | 5,345  (100%) | Sep, 2019 | 162  (5.9%) | 1,236  (45.1%) | 26  (1.0%) | 1,311  (47.8%) | 6  (0.2%) | 2,741  (100%) |
| (13) Insulin lispro | 2,924  (5.3%) | 7,663  (13.8%) | 679  (1.2%) | 44,033  (79.3%) | 212  (0.4%) | 55,511  (100%) | Mar, 2020 | 1,099  (3.7%) | 7,663  (26.0%) | 679  (2.3%) | 19,817  (67.2%) | 212  (0.7%) | 29,470  (100%) |
| Sensitivity analysis** | 2,780  (5.7%) | 8,052  (16.6%) | 483  (1.0%) | 37,059  (76.4%) | 163  (0.3%) | 48,537  (100%) |  | 1,021  (4.0%) | 8,052  (31.5%) | 483  (1.9%) | 15,875  (62.0%) | 163  (0.6%) | 25,594  (100%) |
| (14) Adalimumab | 438  (4.2%) | 1,066  (10.1%) | 40  (0.4%) | 9,002  (85.3%) | 12  (0.1%) | 10,558  (100%) | Jun, 2020 | 219  (4.2%) | 1,066  (20.6%) | 40  (0.8%) | 3,845  (74.2%) | 12  (0.2%) | 5,182  (100%) |
| (15) Insulin aspart | 2,132  (4.3%) | 2,182  (4.4%) | 91  (0.2%) | 45,392  (91.0%) | 62  (0.1%) | 49,859  (100%) | Mar, 2021 | 769  (4.6%) | 2,182  (13.1%) | 91  (0.6%) | 13,561  (81.4%) | 62  (0.4%) | 16,665  (100%) |
| Sensitivity analysis** | 2,090  (4.7%) | 2,241  (5.1%) | 81  (0.2%) | 39,776  (89.9%) | 55  (0.1%) | 44,243  (100%) |  | 758  (5.1%) | 2,241  (15.0%) | 81  (0.5%) | 11,837  (79.1%) | 55  (0.4%) | 14,972  (100%) |
| (16) Ranibizumab | 285  (3.5%) | 1,083  (13.3%) | 5  (0.1%) | 6,784  (83.1%) | 3  (<0.1%) | 8,160  (100%) | Sep, 2021 | 182  (6.0%) | 1,083  (35.5%) | 5  (0.2%) | 1,780  (58.3%) | 3  (0.1%) | 3,053  (100%) |
| (17) Pegfilgrastim | 252  (1.3%) | 269  (1.4%) | 13  (0.1%) | 19,365  (97.2%) | 22  (0.1%) | 19,921  (100%) | Sep, 2023 | 172  (6.7%) | 269  (10.5%) | 13  (0.5%) | 2,090  (81.5%) | 22  (0.9%) | 2,566  (100%) |

Note: For switchers, only the first switch was assessed and counted (i.e., there were some patients who switched twice or more times)

*The first prescription of original biologics and the first prescription of biosimilars occurred in the same month, so that the researchers could not differentiate which was started first.

**While all the original biologics or biosimilars available in Japan were included in the main analysis, the sensitivity analysis made the changes below (corresponding to Supplementary Table S1):

- For somatropin, we considered Genotropin vs. Somatropin BS.
- For erythropoietin, we considered Espo vs. Epoetin Alfa BS (Epoetin Kappa).
- For insulin glargine, we considered Lantus (not including Lantus XR) vs. Insulin Glargine BS.
- For darbepoetin alfa, we considered Nesp vs. Darbepoetin Alfa BS (not including Darbepoetin Alfa authorized generic).
- For insulin lispro, we considered Humalog (not including Humalog Mix and Humalog N) vs. Insulin Lispro BS.
- For insulin aspart, we considered NovoRapid (not including NovoRapid Mix) vs. Insulin Aspart BS.
